# Availability and utilization of new technologies for prevention and control of healthcare-associated infections

**DOI:** 10.1101/2023.03.09.23287055

**Authors:** Pérola Figueiredo Veríssimo, André Ricardo Araujo da Silva

## Abstract

**Background and Aim:** Recently new technologies have emerged to improve prevention and control of healthcare-associated infections. Our aim is to verify availability and use of new technologies for infection controllers.

**Methods:** We conducted a survey in infection controllers from the whole state of Rio de Janeiro state, Brazil, by invitation of a social media group, in August 2022. Nine different technologies were evaluated about availability and use. Categoricals and continuous variables were evaluated by the chi-square test and Mann-Whitney U test, respectively. A value of p less than .05 was considered as statistically significant.

**Results:** One hundred-eight persons answered the questionnaire. The mean age was 42.8 years (24-64 years) and 53/108 (49.1%) reported most workload in public hospitals, 45/108 (41.7%) in private hospitals and 10/108 (9.2%) reported the same workload in public and private hospitals. Out of 108, 63% reported teaching activities in his institutions. There was no correlation between the existence of teaching activities and hospital profile (p=0.42). The most common new technology available was molecular biology (PCR) for microbiological samples research for 73/108 (67.6%) participants. The second new technology available was APP for HAI prevention and control for 33/108 (30.6%), 19/108 (17.6%) reported availability of none technology.

**Conclusion:** Molecular biology (PCR) for microbiological samples research was the most common technology available for infection controllers of an important state of Brazil.

## Introduction

Healthcare-associated infections are one of the most frequent adverse events that occur in acute-care hospitals. These infections cause significant harm to patients and health systems, including the associated increased costs, especially when they are caused by multidrug-resistant organisms. (Amin, 2015). According to the first WHO global report on infection prevention and control launched in 2022, 7% (in high-income countries) and 15% (in low- and middle-income countries) of patients in acute-care hospitals, will acquire at least one HAI during their permanence. (WHO, 2022). According to the Centers for Disease Control and Prevention (CDC), one in 31 hospitalized patients will acquire at least one HAI. (CDC, 2018). Furthermore, according to the European Centre for Disease Prevention and Control, of 142 805 patients staying in an ICU for more than two days (patient-based data), 11 787 (8.3%) patients presented with at least one HAI.

Brazil has been implementing measures to mitigate the occurrence of HAIs. Since 1998, the occurrence of HAI in hospitals has been mandatory for federal law. For this reason, the National Health Surveillance Agency (ANVISA) yearly updates the diagnostic criteria of infection for the epidemiological surveillance of HAIs and the number of them associated with the invasive devices in hospitals. Data of HAIs were obtained on a voluntary basis reported from hospital participants. In the last update of 2021, the global density of incidence of central venous catheter-associated bloodstream infection was 5.16 in adult intensive care units (ICU) and 4.85 in pediatric intensive care units (PICUs). In the same report, the global density of incidence of ventilator-associated pneumonia was 13.1 in ICU and 4,64 in PICUs (ANVISA 2022).

New technological tools can impact HAI control and prevention. (Humphreys, 2010). In this perspective, WHO highlighted the role of new technologies in the field of health, in order to provide support in areas such as training and education. (WHO, 2016). Some of these new technologies aim to influence people ‘s behavior, while others provide direct interference in patient care, such as use of molecular biology to improve identification of multiresistant bacteria, automated cleaning systems, antibiotic-impregnated devices and Apps for infection prevention and control (Blot 2022, Sullivan 2019)

However, further studies need to be carried out to verify the availability of these tools, since they can represent high costs for institutions in a setting of low-middle income countries. (Humphreys, 2010). Considering these aspects our aim is to report availability of nine new technologies for infection controllers of Rio de Janeiro, state, Brazil.

## Methods

We conducted a survey study during August/September 2022 in healthcare workers of Rio de Janeiro state, Brazil that worked as infection control practitioners (IPC). We excluded participants that no resp with incomplete answers

An invitation was sent to a social media group that included professionals from the whole state. After agreement of participants, a questionnaire was sent using google forms. The questions included variables about age, gender, professional category, time (in years) working as IPC, greater workload categorized according to the hospital care (public or private), presence of formal teaching classes for health students in hospitals and type of new technologies used for IPC. We estimated a convened sample of at least 100 participants for analysis.

The data were compiled in an Excel file. We performed a descriptive analysis and used chi-square test for compare categorical data, t test for compare means and Mann–Whitney U test for continuous variables. A value of p less than .05 was considered as statistically significant.

The study was submitted and approved by the Ethics Committee of Faculty of Medicine (Universidade Federal Fluminense), under number 5.563.562 dated from August 4, 2022. A consent statement for the use of data from the each participant included were required.

## Results

One hundred-eight IPC participated in the survey. The mean age of participants was 42,8 years (24-64 years). Demographic data of participants are shown in the table 1

**Table 1.** Demographic data of infection control practitioners of Rio de Janeiro state (Availability and utilization of new technologies for prevention and control of healthcare-associated infections)

| **Variable** | **N (%)** |
| --- | --- |
| **Age (years) N=108** |  |
| 24-30 | 7 (6.5) |
| 31-40 | 43 (40.2) |
| 41-50 | 38 (35.2) |
| 51-60 | 14 (13.1) |
| More than 60 | 6 ( 5.6) |
| **Gender ( N=108)** |  |
| Male | 15 (13.9) |
| Female | 93 (86.1) |
| **Professional category ( N=108)** |  |
| Nurse | 81 (75) |
| Physician | 26 (24.1) |
| Microbiologist | 1 (0.9) |
| **Time working as IPC (in years) N=108** |  |
| Less than 1 | 5 (4.6) |
| 1-5 | 29 (26.9) |
| >5 and <10 | 28 (25.9) |
| >10 and <15 | 25 (23.1) |
| >15 | 21 (19.4) |

Considering the greater workload of IPC according to the hospital type of care, 53/108 (49.1%) worked in public hospitals, while 45/108 (41.7%) in public institutions and 10/108 (9,2%) in both hospitals, with the same workload.

Presence of teaching activities (postgraduate, graduation and specialization programs) in the hospitals of IPC was cited by 68/108 (63%) while 40/108 (37%) informed absence of them.

The availability and utilization of new technologies by IPC are shown in the table 2, according to the specific type Only two (1.9%) participants informed the availability of all technologies. On the other hand, 19/108 (17.6%) reported availability of none technology. At least one technology was available for 25/108 (23.1%) participants, two technologies, at same time, were available and used for 23/108 (21.3%) IPC, three technologies for 17/108 (15.7%) participants and four technologies for 10/108 (9.3%). One participant didn ‘t know if the new technologies were available or not.

**Table 2.** Availability and utilization of new technologies by IPC (Rio de Janeiro state, 2022)

| Technology | Previous utilization (%) | Absence of utilization | Don't know if it's available |
| --- | --- | --- | --- |
| Molecular biology (PCR) for microbiological samples | 73 (67.6) | 32 (29.6) | 3 (2.8) |
| Advanced softwares for HAI analysis | 31 (28.7) | 76 (70.4) | 1 (0.9) |
| Spacial geomapping for HAI identification/ outbreak control | 10 (9.3) | 93 (86.1) | 5 (4.6) |
| Automated decontamination technologies | 15 (13.9) | 92 (85.2) | 1 (0.9) |
| Use of robots for infection control training | 6 (5.6) | 100 (92.6) | 2 (1.9) |
| Ultraviolet devices for environmental cleaning | 31 (28.7) | 76 (70.4) | 1 (0.9) |
| Antibiotic impregnated devices | 22( 20.4) | 84 ( 77.8) | 2 (1.9) |
| Apps for HAI prevention and control | 33 (30.6) | 74 (68.5) | 1 (0.9) |
| Telemedicine | 20 (18.5) | 85 ( 78.7) | 3 (2.8) |

In the table 3, we categorized availability and use of at least one technology, according to the type of assistance and presence or not of teaching activities

**Table 3.**
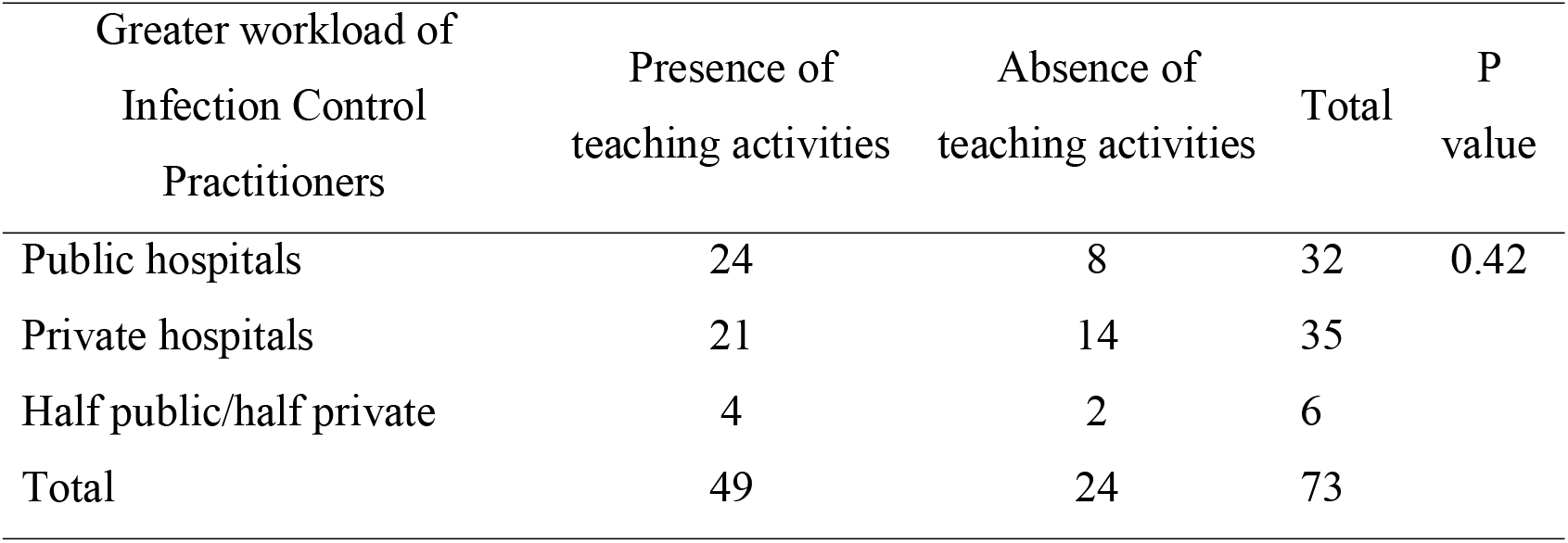
Categorization of greater workload of IPC according to the hospital type of assistance and presence or absence of teaching activities, in units with at least one new technology for infection prevention (Rio de Janeiro state, 2022). N=73

## Discussion

Healthcare-associated infection prevention and control remains a global challenge and since 2016, eight core components for IPC prevention and control were summarized and published by the World Health Organization to be implemented in all countries and healthcare facilities (WHO) (WHO, 2016). Although new technologies were not formally included as core componente, they could be inserted as multimodal strategies in this item (WHO core component 5).

Considering the necessity of research about this topic, in our report, we described availability and use of new technologies for IPC in hospitals from Rio de Janeiro, state, Brazil.

In this survey, most of the responders were nurses, aged between 31 and 50 years, which is similar to found in a large survey conducted in the US and in another study conducted in Santa Catarina, state, Brazil (Pogorzelska 2018, Massaroli 2014). When we evaluated time in years of professionals working as IPC, most of them had more than five years in this area, which resembled the results of US surveys and countries of Middle Eastern (Pororzelska 2018, Tannous 2022).

In Brazil, the healthcare system is based on a public service available for all Brazilian citizens (Brazilian National Health System-SUS) and complementary private services. (Castro 2019) Reflecting this healthcare system, where SUS is the main place of healthcare in Brazil, most of the responders worked as IPC in public services. Interestingly almost 10% of volunteers had the same workload both in public and private services.

We analyzed nine technologies that could help IPC in preventing and avoiding HAI. The most commonly cited available technology was molecular biology (PCR) for microbiological samples identification. Despite molecular biology being not a new technique, its use for prevention and control of HAI is not easy to be implemented in all healthcare facilities (Sader, 1995, Singh-Mooddley 2020). Even with this difficulty, almost two thirds of IPC reported availability to this technology. The second one most cited was use of Apps for HAI prevention and control and although this technology could be easily accessed by all mobiles, quality and relevance of existing Apps could be improved (Bentvelsen 2021).

Other technologies as advanced softwares for HAI analysis, spacial geomapping for HAI identification/ outbreak control, automated decontamination technologies, use of robots for infection control training, ultraviolet devices for environmental cleaning, antibiotic impregnated devices and telemedicine were cited by less than 30% of responders. To our knowledge this manuscript is the first one to describe availability of different technologies to help IPC in HAI prevention, in Brazil.

Simultaneously two or more technologies were available at the same time, to a small proportion of IPC (less than 25%), showing the reality of access to new technologies to prevent HAI in a developing country. In a meeting with 42 experts in infection control prevention, a question about how can technology help to overcome challenges in infection prevention and control (IPC) and to prevent HAI and emerging AMR, four potential domains were identified: 1) role and potential contribution of microbiome research; 2) whole genome sequencing; 3) effectiveness and benefit of antimicrobial environmental surfaces; and 4) future research in hand hygiene. (Zingg 2017). Since then, no new clinical trials have identified the true impact of new technologies used at the same time to prevent IPC in a “real world”.

As expected, new technologies were available more frequently in hospitals (both public and private) with teaching activities when compared with hospitals with absence of teaching activities, although without statistical significance. Presence of teaching activities in hospitals help to improve infection control practices and adherence to protocols in order to avoid infections (Sekimoto 2008)

## Conclusion

Molecular biology (PCR) for microbiological samples research was the most common technology available for infection controllers in a middle-income state of Brazil, available for almost three quarters of participants.

## Data Availability

All data produced in the present study are available upon reasonable request to the authors

## Notes

### Competing Interest Statement

The authors have declared no competing interest.

### Funding Statement

This study did not receive any funding

## References

1- Amin AN, Deruelle D. Healthcare-associated infections, infection control and the potential of new antibiotics in development in the USA. Future Microbiol. 2015 Jun;10(6):1049–62. doi: 10.2217/fmb.15.33.

2- WHO launches first ever global report on infection prevention and control. World Health Organization, Geneve, 2022. Available from https://www.who.int/news/item/06-05-2022-who-launches-first-ever-global-report-on-infection-prevention-and-control. Accessed on February 16, 2023.

3- Healthcare-associated infections acquired in intensive care units Annual Epidemiological Report for 2017. European Centre for Disease Prevention and Control. Available from https://www.ecdc.europa.eu/sites/default/files/documents/AER_for_2017-HAI.pdf Accessed on February 16, 2023.

4- Boletim Segurança do Paciente e Qualidade em Serviços de Saúde nº 28-Agência Nacional de Vigilância Sanitária-Brasil, 2022.. Available from https://app.powerbi.com/view?r=eyJrIjoiZDIwZjYyMzUtMmYxZS00MTRjLTk0NWMtZWE2ZDUzOGRjOTVjIiwidCI6ImI2N2FmMjNmLWMzZjMtNGQzNS04MGM3LWI3MDg1ZjVlZGQ4MSJ9. Accessed on February 16, 2023.

5- Humphreys H. New technologies in the prevention and control of healthcare-associated infection. J R Coll Physicians Edinb. 2010 Jun;40(2):161–4.

6- World Health Organization. Guidelines on core components of infection prevention and control programmes at the national and acute health care facility level. Geneva: WHO; 2016. Available at: https://www.who.int/gpsc/core-components.pdf. Access on September 25, 2022.

7- Blot S, Ruppé E, Harbarth S, Asehnoune K, Poulakou G, Luyt CE, Rello J, Klompas M, Depuydt P, Eckmann C, Martin-Loeches I, Povoa P, Bouadma L, Timsit JF, Zahar JR. Healthcare-associated infections in adult intensive care unit patients: Changes in epidemiology, diagnosis, prevention and contributions of new technologies. Intensive Crit Care Nurs. 2022 Jun;70:103227. doi: 10.1016/j.iccn.2022.103227.

8- Sullivan KV, Dien Bard J. New and novel rapid diagnostics that are impacting infection prevention and antimicrobial stewardship. Curr Opin Infect Dis. 2019 Aug;32(4):356–364

9- Pogorzelska-Maziarz M, Gilmartin H, Reese S. Infection prevention staffing and resources in U.S. acute care hospitals: Results from the APIC MegaSurvey. Am J Infect Control. 2018 Aug;46(8):852–857. doi: 10.1016/j.ajic.2018.04.202. Epub 2018 Jun 1. PMID: 29861151.

10- Massaroli A, Martini JG. PERFIL DOS PROFISSIONAIS DO CONTROLE DE INFECÇÕES NO AMBIENTE HOSPITALAR. Cienc Cuid Saude 2014 Jul/Set; 13(3):511–518

11- Tannous E, El-Saed A, Ameer K, Khalaf A, Mohammad S, Molaeb B, Alshamrani MM. Infection prevention and control staffing and programs in Middle Eastern Countries. J Infect Dev Ctries. 2022 May 30;16(5):889–896. doi: 10.3855/jidc.15504. PMID: 35656962.

12- Castro MC, Massuda A, Almeida G, Menezes-Filho NA, Andrade MV, de Souza Noronha KVM, Rocha R, Macinko J, Hone T, Tasca R, Giovanella L, Malik AM, Werneck H, Fachini LA, Atun R. Brazil ‘s unified health system: the first 30 years and prospects for the future. Lancet. 2019 Jul 27;394(10195):345–356. doi: 10.1016/S0140-6736(19)31243-7. Epub 2019 Jul 11. PMID: 31303318.

13- Sader HS, Hollis RJ, Pfaller MA. The use of molecular techniques in the epidemiology and control of infectious diseases. Clin Lab Med. 1995 Jun;15(2):407–31. PMID: 7671580.

14- Singh-Moodley A, Ismail H, Perovic O. Molecular diagnostics in South Africa and challenges in the establishment of a molecular laboratory in developing countries. J Infect Dev Ctries. 2020 Mar 31;14(3):236–243. doi: 10.3855/jidc.11779. PMID: 32235082.

15- Bentvelsen RG, Holten E, Chavannes NH, Veldkamp KE. eHealth for the prevention of healthcare-associated infections: a scoping review. J Hosp Infect. 2021 Jul;113:96–103. doi: 10.1016/j.jhin.2021.04.029. Epub 2021 May 3. PMID: 33957179.

16- Zingg W, Park BJ, Storr J, Ahmad R, Tarrant C, Castro-Sanchez E, Perencevich E, Widmer A, Krause KH, Kilpatrick C, Tomczyk S, Allegranzi B, Cardo D, Pittet D; 2017 Geneva IPC-Think Tank. Technology for the prevention of antimicrobial resistance and healthcare-associated infections; 2017 Geneva IPC-Think Tank (Part 2). Antimicrob Resist Infect Control. 2019 May 22;8:83. doi: 10.1186/s13756-019-0538-y. PMID: 31139366; PMCID: PMC6530187.

17- Sekimoto M, Imanaka Y, Kobayashi H, Okubo T, Kizu J, Kobuse H, Mihara H, Tsuji N, Yamaguchi A; Japan Council for Quality Health Care, Expert Group on Healthcare-Associated Infection Control and Prevention. Impact of hospital accreditation on infection control programs in teaching hospitals in Japan. Am J Infect Control. 2008 Apr;36(3):212–9. doi: 10.1016/j.ajic.2007.04.276. PMID: 18371518.

